# Interpreting Wastewater SARS-CoV-2 Results using Bayesian Analysis

**DOI:** 10.1101/2021.08.17.21262165

**Authors:** Kyle Curtis, Raul A. Gonzalez

## Abstract

Wastewater surveillance of severe acute respiratory syndrome coronavirus 2 (SARS-CoV-2) has proven a practical complement to clinical data for assessing community-scale infection trends. Clinical assays, such as the CDC-promulgated N1, N2, and N3 have been used to detect and quantify viral RNA in wastewater but, to date, have not included estimates of reliability of true positive or true negative. Bayes’ Theorem was applied to estimate Type I and Type II error rates for detections of the virus in wastewater. Conditional probabilities of true positive or true negative were investigated when one assay was used, or multiple assays were run concurrently. Cumulative probability analysis was used to assess the likelihood of true SARS-CoV-2 detection using multiple samples. Results demonstrate highly reliable positive (>0.86 for priors >0.25) and negative (>0.80 for priors = 0.50) results using a single assay. Using N1 and N2 concurrently caused greater reliability (>0.99 for priors <0.05) when results concurred but generated potentially counterintuitive interpretations when results were discordant. Regional wastewater surveillance data was investigated as a means of setting prior probabilities. Probability of true detection with a single marker was investigated using cumulative probability across all combinations of positive and negative results for a set of three samples. Findings using a low (0.11) and uniformed (0.50) initial prior resulted in high probabilities of detection (>0.95) even when a set of samples included one or two negative results, demonstrating the influence of high sensitivity and specificity values. Analyses presented here provide a practical framework for understanding analytical results generated by wastewater surveillance programs.

## Introduction

The on-going coronavirus disease-2019 (COVID-19) pandemic has reached most regions of the world, with global cases and deaths currently greater than 185 million and 4 million, respectively (Dong et al., 2020). COVID-19 wastewater surveillance has emerged as a complementary epidemiological tool used to identify community level infection trends, as an early warning system, and even to screen for infected individuals at the building level. The dominant analytical techniques to quantify severe acute respiratory syndrome 2 (SARS-CoV-2), the virus that causes COVID-19, are PCR-based (e.g., RT-qPCR, RT-dPCR) and use clinical assays. Globally, the research community (Bivins et al., 2020; Naughton et al. 2021) has made considerable progress in effectively applying this tool to quantify the virus in wastewater and observe community-scale trends. The next phase of implementation should focus on appropriate interpretation of results with respect to confidence or probability.

Bayes’ Theorem can be used to calculate results in terms of likelihood, or in the language of Bayesian statistics, posterior probability. Because of type I and II errors in PCR data, results are more accurately reported in terms of probability, which includes a consideration of prior likelihood of an event as well as false positive and negative rates. This approach computes the conditional probability of reliable result (true negative or true positive), then continually updates prior assumptions as new information is acquired. A Bayesian framework for interpreting analytical results from environmental samples has been applied in the field of microbial source tracking (MST) (Kildare et al. 2007; Johnston et al. 2013, Curtis and Gonzalez 2019, Chen et al. 2021). Here PCR markers for human-associated fecal microbes are used to identify waterways potentially impacted by wastewater. Employing a Bayesian approach characterizes results in terms of contamination probability rather than simple presence/absence or concentration of the marker. This method provides a quantitative way to incorporate new information (e.g. recent sample results) into the interpretation of subsequent results.

Applying a probabilistic framework for understanding the reliability of results requires only the sensitivity and specificity of the analytical test and the ability to set a prior. Here we examine this approach using the Centers for Disease Control (CDC) SARS-CoV-2 panel (N1, N2, N3) under a variety of wastewater surveillance scenarios. The likelihood of true positive and true negative is examined using individual assays across a range of prior probabilities and also using multiple assays concurrently. A proposed technique for setting realistic priors based on wastewater sampling results is described using data from a wastewater surveillance network with nine treatment facilities in southeastern Virginia. Finally, cumulative probability is used to investigate the interpretation of multiple sample results under two scenarios.

## Methods

### CDC Assay Sensitivity and Specificity

The CDC SARS-CoV-2 real-time reverse transcription PCR panel (N1, N2, N3) was used to assess the likelihood that wastewater samples contained SARS-CoV-2 RNA based on analytical results, and assay sensitivity and specificity. These assays were developed for clinical identification of SARS-CoV-2 infection but have been widely applied to environmental samples, particularly wastewater, for virus detection and enumeration (e.g. Stadler et al. 2020, Agrawal et al. 2021, McClary-Gutierrez 2021). The specificity of each assay was tested in-vitro by Lu et al. (2020) finding N1, N2 and N3 assays with specificity values of 1.0, 1.0, and 0.955 respectively. Specificity can be described as the tendency of an assay to not detect the target of interest when it is absent and is calculated by dividing the number of true negative values by the number of test samples which did not contain the target. Thus, a specificity of 0.955 suggests a false-negative rate of 4.5%.

The sensitivity of an assay describes its ability to detect the target when it is present. Wastewater sensitivity values were calculated using regional clinical data from the Virginia Department of Health (https://www.vdh.virginia.gov/coronavirus/) and SARS-CoV-2 wastewater results (Gonzalez et al. 2020). Clinical PCR positive case counts by zip code for all zip codes within the 9 wastewater treatment facility sewersheds in Hampton Roads, Virginia were gathered from May 15, 2020 through November 17, 2020. True positives were defined as when a weekly wastewater sample from a facility was positive and at least one zip code in the facility’s catchment had a clinically confirmed infected individual that week. For each of the 3 assays, the number of weekly wastewater samples from the 9 facilities were pooled. The number of samples with a true positive were divided by the total number of samples to determine the sensitivity of each assay in wastewater. The sensitivity for N1, N2, and N3 assays are 75% (200/266), 83% (220/266), and 80% (212/266) respectively. Sensitivity and specificity as used in subsequent equations are described below.

Sensitivity (Se) and Specificity (Sp);

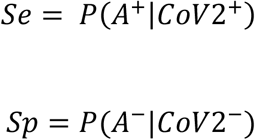

where;

Se = assay sensitivity

Sp = assay specificity

A^+^ = a positive detection with a given assay

A^-^ = a negative analytical result with a given assay

### Bayes’ Theorem

Bayes’ Theorem (eq 1) was applied to CDC assay sensitivity and the wastewater-derived specificity to investigate the likelihood that SARS-CoV-2 is present in wastewater samples given positive and negative analytical results.

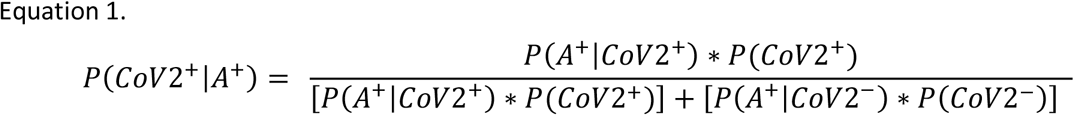

where;

CoV2^+^ = SARS-CoV-2 is present

CoV2^-^ = SARS-CoV-2 is absent

The posterior probability that SARS-CoV-2 is present in a sample given detection of each assay (N1, N2, N3) was tested across all possible priors (0.0-1.0) using Equation 1. A similar conditional probability was derived to test the likelihood that SARS-CoV-2 is absent from a sample given a negative assay result and was also tested across all priors.

As noted, the specificity of the N1 and N2 assays determined by Lu et al. were 1.0. A specificity value of 1.0 guarantees the absence of false positive results regardless of assay sensitivity and prior probability. As a result, it is difficult to observe the effect of differing approaches for setting priors and the implications of using one or multiple assays. To investigate these factors, we have used a specificity value of 0.999 for all calculations.

### Surveillance Using Multiple Assays Concurrently

A multiple-assay approach to environmental sampling has been supported in the field of microbial source tracking (MST) (Harwood et al. 2014; Ahmed et al. 2016; Jardé et al. 2018;) and is currently employed by many researchers conducting wastewater surveillance of SARS-CoV-2 (Stadler et al. 2020; Peñarrubia et al. 2020; Agrawal et al. 2021). The likelihood that SARS-CoV-2 is present in a sample was investigated using multiple assays concurrently. Equation 2 describes the probability that SARS-CoV-2 is present given positive detection of the virus with two assays. N1 and N2 assay were selected for this analysis due to CDC’s recommendation to remove N3 from the clinical panel (Lu et al. 2020). An analogous equation was used to test the likelihood that SARS-CoV-2 is present when failing to detect the virus using two assays. All combinations of N1 and N2 detection and non-detection were tested across the range of possible priors (0.0-1.0).

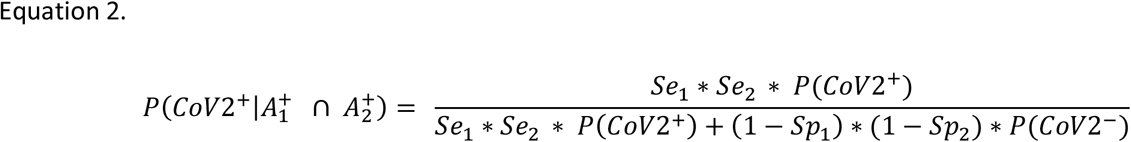

where;

Se_i_ = sensitivity of assay *i*

Sp_i_ = specificity of assay *i*

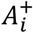 = detection of assay *i*

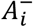 = failing to detect assay *i*

### Prior Probabilities Based on Wastewater Detection

Wastewater surveillance data using the N2 assay and the N1 and N2 assays concurrently were used to set priors based on percent positive wastewater samples. In this case the prior probability of SARS-CoV-2 in wastewater was set as the percent of samples which were positive for one or both assay in the week prior. Wastewater data for these analyses come from the wastewater surveillance program being conducted by the Hampton Roads Sanitation District (HRSD) and comprise a raw influent sample from each of 9 regional treatment facilities. For a detailed description of sampling and analytical methodologies see Gonzalez et al. 2020. Using wastewater-derived priors, posterior probability of SARS-CoV-2 presence was calculated for wastewater samples using N2 alone and N1 and N2 concurrently. For samples that were negative for N2 or both N1 and N2 the posterior probability that SARS-CoV-2 is absent in a given wastewater sample was reported.

### Cumulative Probability from Multiple Samples

The cumulative probability of SARS-CoV-2 based on results from three separate samples was investigated for two scenarios representing a comparatively low and high initial prior. The low initial prior (0.11) was based on the lowest possible non-zero value for percent positive treatment facilities (1 of 9 facilities with a detection) in the Hampton Roads wastewater surveillance network. The high initial prior (0.50) was used to examine a situation where there is no reliable way to estimate prior likelihood and thus priors are uninformed. Cumulative probability of the presence of SARS-CoV-2 was calculated using Equation 4 for each sample and the resulting posterior probability was used as the prior for subsequent samples.

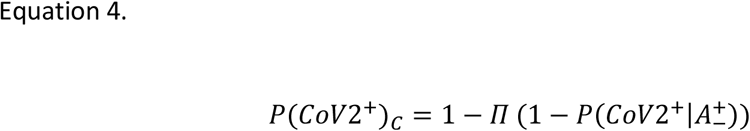

Where;

P(CoV2^+^)_c_ = cumulative probability of SARS-CoV-2

P(CoV2^+^|A±) = probability of SARS-CoV-2 given a positive or negative assay

## Results and Discussion

### Confidence Using a Single Assay

Bayes’ Theorem was applied in conjunction with sensitivity and specificity values for the three CDC assays to calculate posterior probabilities of true positive and true negative across all possible priors (Figure 1). The high specificity values of each assay resulted in expectedly high posterior probabilities of SARS-CoV-2 given a positive analytical result (Figure 1a). The resulting curves suggest that when N1 or N2 are used, a positive analytical result is highly reliable (>0.97), even at priors below 0.05. While not as overwhelming as N1 or N2, the N3 assay also generated highly reliable positive results with posterior probabilities of SARS-CoV-2 >0.86 for priors >0.25. The marginally increased likelihood of false positive results for N3 was expected given the slightly lower specificity (0.955). As described by Lu et al., the N3 assay was intentionally designed to also detect SARS-CoV, resulting in a lower specificity value when cross-reactivity was tested against closely related respiratory viruses. The likelihood of obtaining a true negative result was high with all assays when SARS-CoV-2 was absent (Figure 1b). Values denoting likelihood of true negative result are more similar among assays than those for true positives, owing to the similarity in calculated sensitivity values. Findings suggest that when using an uninformed prior (0.50) a single assay generates acceptable confidence (0.80) in negative results. Given no additional data to inform prior selection all assays performed well and result in a reasonably high likelihood of true negative results. If there is reason to reduce the estimate of prior probability the reliability of a true negative increases dramatically and is greater than 0.92 for all priors <0.25 (Figure 1).

**Figure 1.**
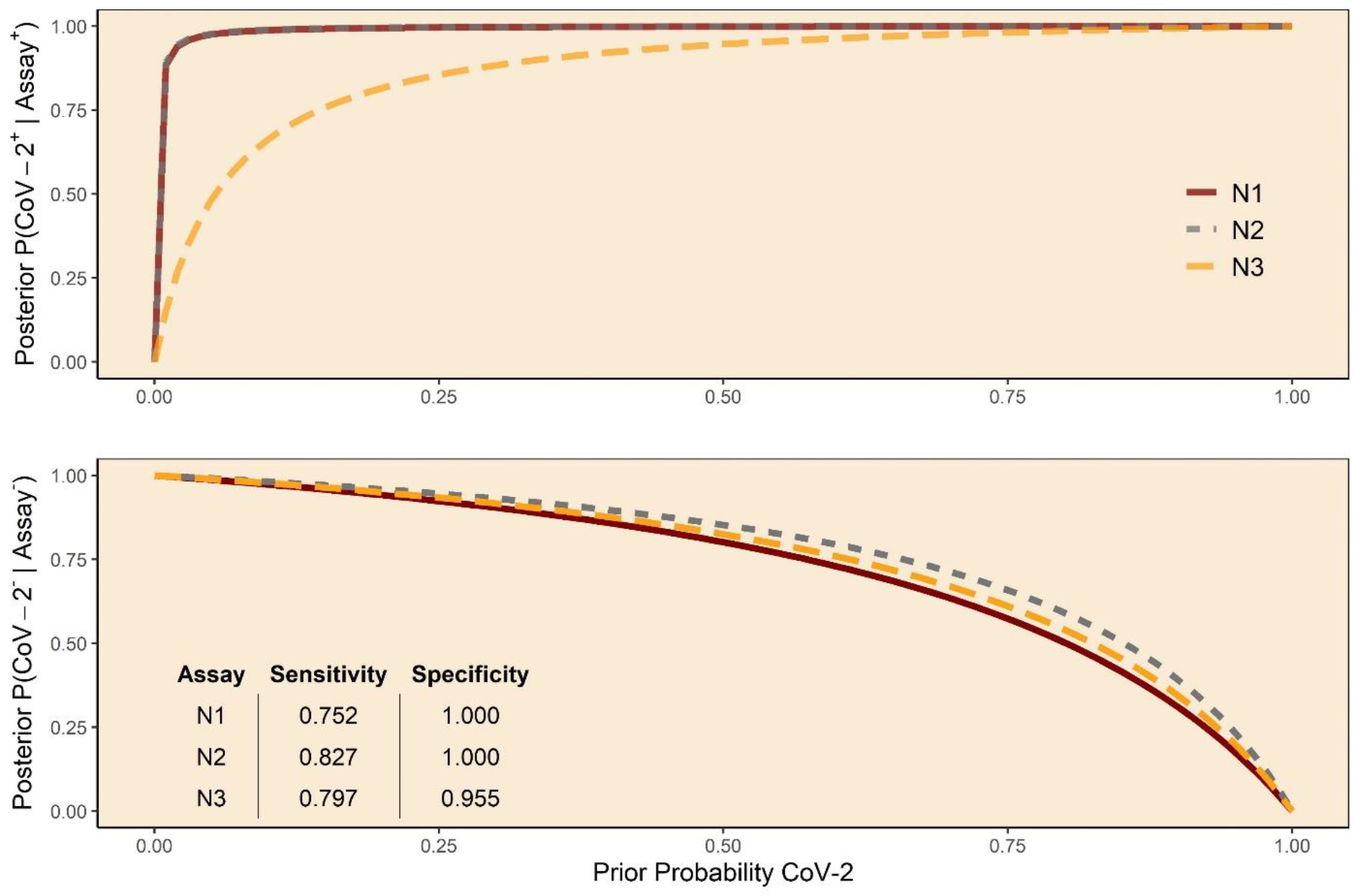
Posterior probability of true positive (top) and true negative (bottom) result for the N1, N2, and N3 assays across all possible prior probabilities.

### Surveillance Using Multiple Assays Concurrently

Many COVID-19 wastewater surveillance publications have concurrently used multiple SARS-CoV-2 assays. When different combinations of N1 and N2 assay results were tested across all priors, the resulting posterior probabilities for SARS-CoV-2 presence track in a generally predictable manner (Figure 2). When both assays detect the virus there is an exceedingly high probability (>0.99) that this detection is reliable, even at very low priors (0.005). Similarly, when either assay is used individually and detects the virus, these results are also highly reliable, as described in the previous section. Discordant results (N1+/N2- or N2+/N1-) using two assays also yields a high probability that SARS-CoV-2 is present (>0.87), even when priors are low (0.05). This is a potentially counterintuitive result which requires an understanding of test characteristics (sensitivity and specificity) as well as the effect of prior probability to appropriately interpret. Failing to consider these results in a Bayesian framework, one might tacitly assign a likelihood of 0.5 to the probability that SARS-CoV-2 is present, given that two highly accurate tests yielded conflicting results. Such an interpretation not only underestimates the likelihood that SARS-CoV-2 is present, it also provides no more information about SARS-CoV-2 presence in wastewater than what was known prior to sampling. In other words, this interpretation suggests that the virus is just as likely present as not. Alternatively, one might assume a low prior probability that SARS-CoV-2 is present, given low community incidence at the time of sampling, and therefore conclude that conflicting results actually suggest the virus is absent by considering the one positive result a false positive. This would also be an incorrect interpretation, given what is known about the sensitivity and specificity of these assays. The implication of this interpretation is assigning a 0.0 probability that SARS-CoV-2 is present when in fact there is a >0.93 likelihood that it is present for all priors above 0.10.

**Figure 2.**
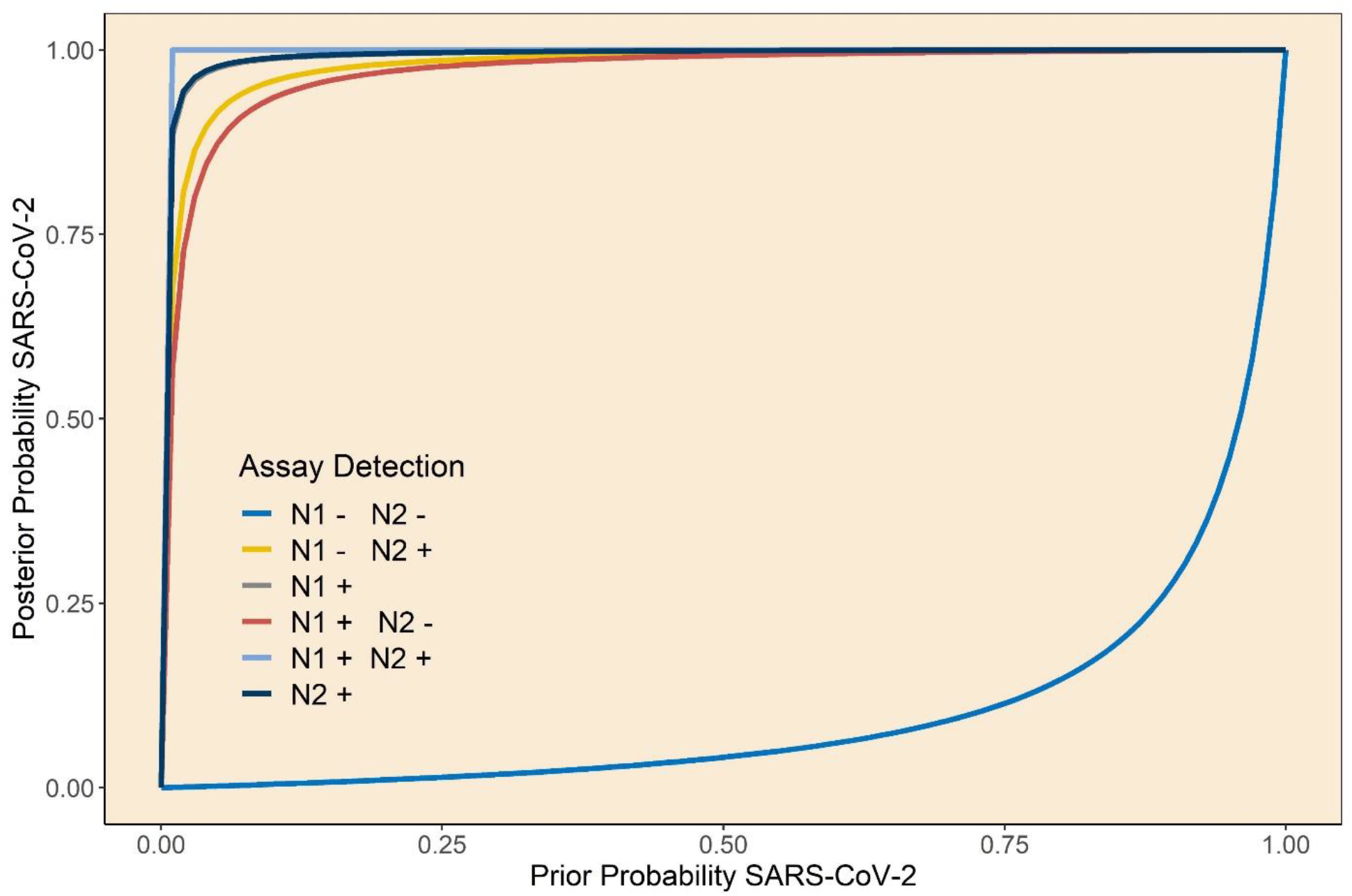
Posterior probability that SARS-CoV-2 nucleic acid is present given combinations of results using one or both of the N1 and N2 assays. Posteriors plotted across all possible prior probabilities that SARS-CoV-2 is present.

### Wastewater Detections to Inform Priors

One strategy for selecting prior probability of SARS-CoV-2 in wastewater samples is to use recent wastewater surveillance results. Surveillance programs often sample at routine intervals, and some include multiple facilities. Therefore, a simple metric such as percent positive samples over recent weeks or across multiple facilities can be used to inform prior probability. Relatively large priors (0.39-1.0) resulted when percent positive facilities were calculated using the Hampton Roads surveillance data due to the ubiquity of the virus in this region for the study period. With most priors being >0.50 positive results are highly reliable for all samples, regardless if one or both assays were used (Fig 3). These findings suggest that a single assay likely provides sufficient confidence in positive results when detections are routinely seen at monitored facilities, causing priors to be set relatively high.

**Figure 3.**
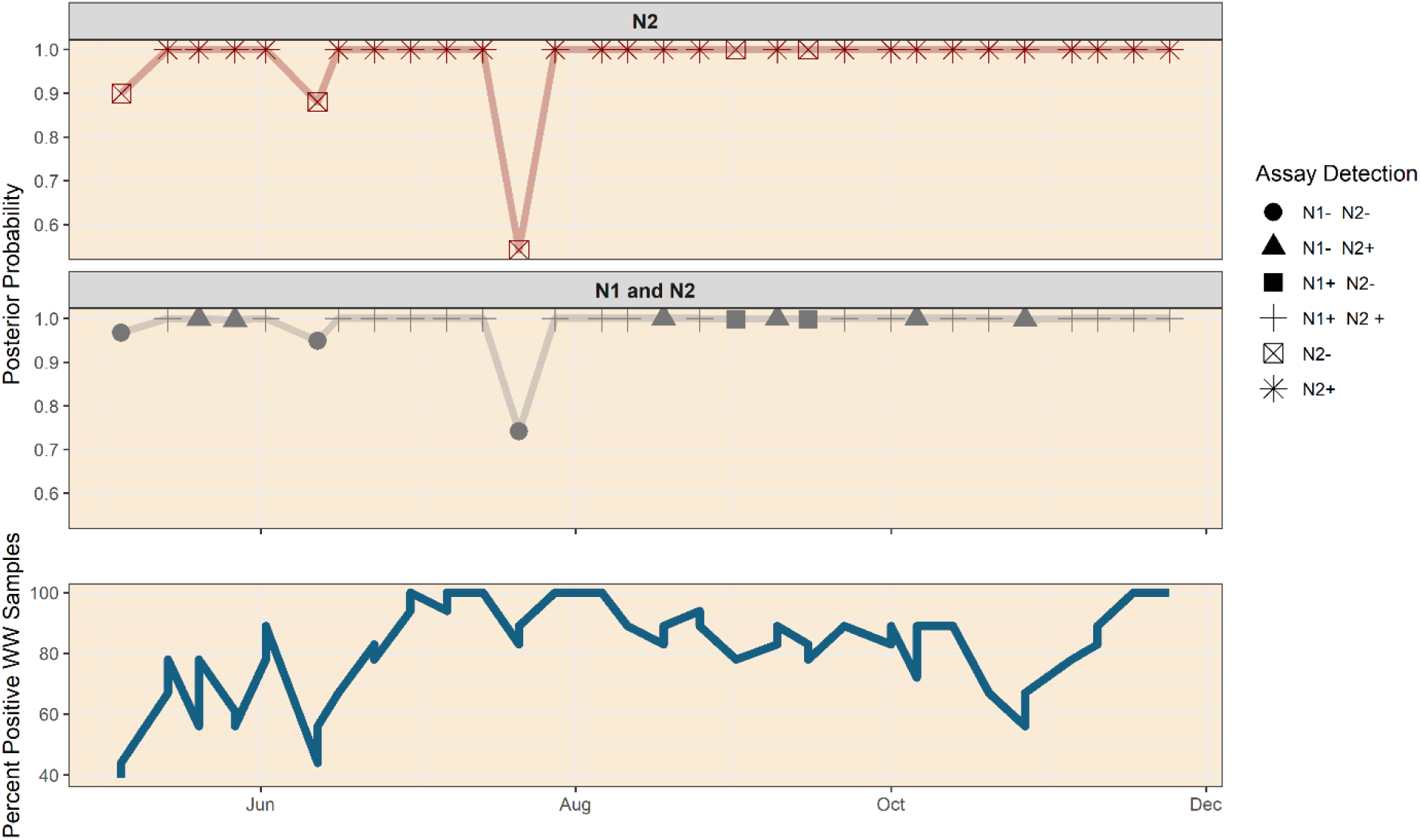
Posterior probability of true positive or true negative using the N2 assay (top). The middle panel shows the likelihood of true positive given detection with one or both of N1 and N2 assays and true negative when both assays used concurrently fail to detect the virus. The bottom panel indicates the number of treatment facilities (n=9) in which the virus was detected. These values are used as priors for calculating posterior probabilities plotted in the top and middle panels.

**Figure 4.**
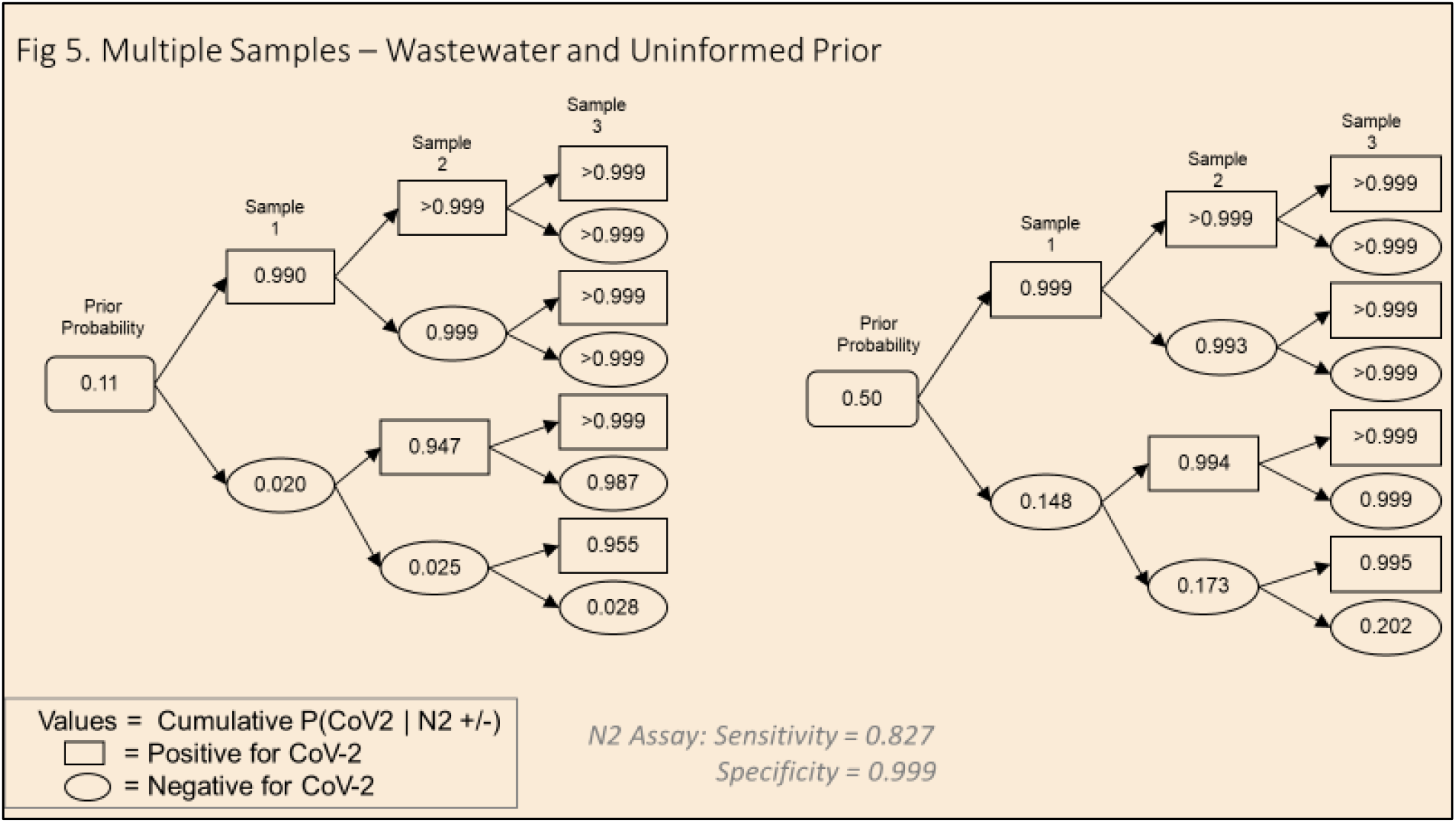
Cumulative probability that SARS-CoV-2 nucleic acid is present given two initial priors (0.11, 0.50). Cumulative posteriors are plotted across three sampling events with all combinations of positive/negative results displayed.

Conversely, the likelihood of true negative is somewhat driven down (Figure 3) when wastewater-derived priors result in high values. This effect is understandable considering that numerous recent detections should suggest a greater likelihood of detection for the current set of samples. The consequence of high priors in constraining reliability of negative results is clearly shown in the 7/21/20 data from Figure 3. This figure shows the likelihood of true positive for samples given detection of one or both assay and likelihood of true negative when one or both assay<u>s</u> fail to detect viral RNA. Here, given a prior probability of 83% that SARS-CoV-2 is present, a single negative N2 result indicates a 54% likelihood of true negative. Finding negative results for both assays increases the likelihood of true negative, but only to 74%. This is a significant departure for the reliability associated with concordant negative results when priors are lower (<0.15), which approaches 100%. In a scenario of high priors based on numerous recent detections a negative result should be assigned less reliability than one when few recent samples are positive, given that wastewater detection of viral RNA is an indication of community incidence for a catchment. However, such an interpretation may not be intuitive if one simply considers the sensitivity of the assay but fails to account for the prior likelihood of detection.

Finding conflicting results when running multiple assays in the context of higher, wastewater-derived priors also produces potentially difficult to interpret results. For the range of wastewater-derived priors tested (0.39-1.0), discordant results regardless of which assay was detected, provide strong evidence that SARS-CoV-2 is present (Figure 3). As described above, failing to consider prior probability of detection could lead a researcher to interpret conflicting results as a 0.5 likelihood that SAR-CoV-2 RNA is present in the sample. Interpretation using a Bayesian framework would suggest that, even at the lowest prior for which there were conflicting results (0.56), the likelihood that the virus is present is >0.99.

### Multiple Samples

Collecting multiple samples is an effective alternative for increasing reliability in results while avoiding the need to run multiple assays. This approach generates high confidence via the cumulative probability of true positive/negative results across samples, rather than the intersection of positive/negative results for two or more assays. A multiple sample strategy was applied using a prior of 0.11 to examine the effect of a wastewater-derived prior for a situation where one of the nine facilities in the HRSD surveillance network had a positive detection in the previous week. At this prior, any combinations of samples that include a positive result yield >0.95 likelihood that viral RNA is present. A negative result in sample one reduces the likelihood that SARS-CoV-2 is present from 0.11 to 0.02. In this scenario the second sample is highly informative. Finding a positive result in sample two, even after a negative for sample one, yields a 0.95 likelihood that the virus is present. High cumulative probability that the virus is present given conflicting results from two samples using a single assay is consistent with the analogous finding of high likelihood given the intersection of two conflicting, different assays run on a single sample. With no detection in sample one, a second negative result has little impact on the cumulative probability, which is 0.025, giving good evidence that the virus is absent. This pattern continues for each sample three endpoint, wherein any positive result drives the cumulative probability that SARS-CoV-2 is present in a set of three samples above 0.94 while consecutive negative results yield cumulative probabilities (0.020 – 0.028) which remain one order of magnitude lower than the prior (0.11).

Using an uniformed prior (0.50) results in a similarly high posterior probability (>0.99) that SARS-CoV-2 is present for all combinations of sampling results other than three consecutive negatives. A key difference with this prior is that even samples with only negative results still yield a 0.15-0.20 likelihood that SARS-CoV-2 is present. As described previously, this strategy for setting priors is reasonable when there is no reliable way to estimate likelihood of detecting viral RNA before samples have been analyzed. Using a prior of 0.50 suggests a 50% likelihood that a sample contains the SARS-CoV-2 virus at a concentration greater than the analytical limit of detection. The interaction of a prior that suggests an equal chance of detecting RNA as not (0.50) with the high sensitivity and specificity of the N2 assay generates some cumulative probabilities that may seem counterintuitive. The main examples include sets of three samples for which two results are negative and one is positive but the cumulative probability that SARS-CoV-2 is present is still >0.99. Intuitively, two negative results, regardless of where they occur in the sequence of samples, seem to suggest the absence of SARS-CoV-2. Following a Bayesian approach in which priors are updated each time new data is acquired helps clarify these results. As described, the posterior probability from each sample is used as the prior for subsequent samples. Sets of samples where the positive result occurred first in sequence results in high posterior probabilities which are carried through as priors for the remaining samples. For these cases, even negative downstream results yield high posterior probabilities, driven by priors of >0.99 used in samples two and three. In cases where sample one is negative, the prior of 0.50 and high sensitivity and specificity prevent the posterior probability from being driven below 0.14. As a result, a positive detection in sample one or two results in a cumulative probability of >0.99.

### Limitations and Assumptions

Applying a Bayesian framework for interpreting analytical findings is fundamentally driven by the sensitivity and specificity values assigned to the assays in question, along with the assumed prior. As a result, the accuracy of these values is highly consequential for the analysis. For this preliminary exercise, in vivo specificity values similar to those reported by Lu et al. (2020) were used in conjunction with a novel in situ approach to determining sensitivity values from a regional SARS-CoV-2 wastewater monitoring program. As noted, specificity values of 0.999 were used in place of the 1.0 specificities reported by Lu et al. (2020) for the N1 and N2 assays. The goal of this approach was to assume a false positive rate of 1/1000 in order to conservatively examine the reliability of analytical results when neither sensitivity nor specificity are perfect. Where possible, future studies should conduct in-house specificity testing using their own analytical methodology performed on the matrix being studied to derive lab-specific specificity values. A similar recommendation is suggested for deriving lab-specific sensitivity values and is likely more impacted by wastewater matrix effects such as inhibition.

After establishing assay sensitivity and specificity, prior probability is the final value required to conduct the simple conditional probability analyses described above. Prior probability is another critical piece of this framework as it also exerts a significant influence on the outcome and the process for setting a value may not be intuitive. Establishing the prior probability of an event or result can be challenging when little is known about the system being studied. Regarding wastewater-based epidemiology one suggested approach for estimating prior probability of SARS-CoV-2 detection is to use recent results from the same location (e.g., the proportion of positive results from the last set or sets of samples) or results from other areas within a common region. Priors may also comprise a distribution, allowing for some incorporation of uncertainty. For instance, noting that clinical cases in the area of study have been increasing rapidly in the preceding weeks does not suggest a specific value for use as a prior probability but does provide information which should not be ignored when assessing reliability of wastewater detections. Integrating this clinical information into a Bayesian analysis of wastewater results might include setting priors using a uniform distribution of 0.51-0.99, implying that little is known about the true prior probability of SARS-CoV-2 detection, but regional case data suggests that a detection is likelier than not. Performed this way the posterior probability is also a distribution which allows for uncertainty around the estimate to be easily included.

## Data Availability

Data referred to in this manuscript can be made available upon request.

## AUTHOR INFORMATION

### Author Contributions

The manuscript was written through contributions of all authors. All authors have given approval to the final version of the manuscript.

